# Stage-wise algorithmic bias, its reporting, and relation to classical systematic review biases in AI-based automated screening in health sciences: A structured literature review

**DOI:** 10.1101/2025.05.16.25327774

**Authors:** José Luis Pardal-Refoyo, Beatriz Pardal-Pelaéz

**Affiliations:** University Hospital of Salamanca. Otorhinolaryngology and Head and Neck Surgery. University of Salamanca. Department of Surgery. Faculty of Medicine. Otorhinolaryngology Area. IBSAL. Salamanca. Spain; University of Salamanca. Faculty of Medicine and Dental Clinic. Department of Surgery. Stomatological Area. Salamanca. Spain

**Keywords:** Algorithmic bias, Artificial intelligence, Systematic Review, Health Sciences, Equity, Reporting Standards

## Abstract

**Introduction:** Algorithmic bias in systematic reviews that use automatic screening is a major challenge in the application of AI in health sciences. This article presents preliminary findings from the project titled “Identification, Reporting, and Mitigation of Algorithmic Bias in Systematic Reviews with AI-Assisted Screening: Systematic Review and Development of a Checklist for its Evaluation” registered in PROSPERO with the registration number CRD420251036600 (https://www.crd.york.ac.uk/PROSPERO/view/CRD420251036600). The results presented here are preliminary and part of ongoing work.

**Objective:** To synthesize knowledge about the taxonomies of algorithmic bias, reporting, relationships with classical biases, and use of visualizations in AI-supported systematic reviews in health sciences.

**Methods:** A specific literature review was conducted, focusing on systematic reviews, conceptual frameworks, and reporting standards for bias in AI in healthcare, as well as studies cataloguing detection and mitigation strategies, with an emphasis on taxonomies, transparency practices, and visual/illustrative tools.

**Results:** A mature body of work describes stage-based taxonomies and mitigation methods for algorithmic bias in general clinical AI. Common improvements in reporting and transparency (e.g. CONSORT-AI, SPIRIT-AI) are described. However, there is a notable absence of direct application to AI-automated screening of systematic reviews or empirical analyses of the interactions of biases with classical biases at the review level. Visualization techniques, such as bias heatmaps and pipe diagrams, are available, but have not been adapted to review workflows.

**Conclusions:** There are fundamental methodologies to identify and mitigate algorithmic bias in AI in health, but significant gaps remain in the understanding and operationalization of these frameworks within AI-assisted systematic reviews. Future research should address this translational gap to ensure transparency, fairness, and methodological rigor in the synthesis of evidence.

## Introduction

Algorithmic bias has become a critical concern in the expansion of artificial intelligence (AI) in healthcare applications, where systematic errors can arise at multiple stages of model development and deployment, leading to inequitable outcomes for diverse populations ^1–3^. The proliferation of AI tools in the life sciences requires robust frameworks for the identification, reporting, and mitigation of bias ^1,2,4^. A bias of comprehensive taxonomies mapping each phase of the machine learning lifecycle has been developed in recent years, as well as the proposal of enhanced reporting standards (e.g., CONSORT-AI, SPIRIT-AI) to ensure transparency and accountability ^2,4^. Despite these advances, the adaptation and evaluation of these frameworks within AI-backed evidence synthesis, specifically, systematic reviews incorporating automated study selection remain unexplored in the current literature.

The aim is to develop graded taxonomies of algorithmic biases, their reporting mechanisms, and interactions with classic review biases in the context of AI-based screening in health sciences, including relevant visualizations and interdisciplinary examples.

This article presents preliminary findings from the project titled “Identification, Reporting, and Mitigation of Algorithmic Bias in Systematic Reviews with AI-Assisted Screening: Systematic Review and Development of a Checklist for its Evaluation” registered in PROSPERO with the registration number CRD420251036600 (https://www.crd.york.ac.uk/PROSPERO/view/CRD420251036600). The results presented here are preliminary and part of ongoing work.

## Methods

Overview of the available literature on algorithmic bias taxonomies, identification and mitigation strategies, and reporting practices relevant to the health sciences, with particular attention to their application (or lack thereof) in systematic reviews using AI-based automated screening. The databases consulted for the identification and selection of relevant literature included: PubMed (https://pubmed.ncbi.nlm.nih.gov/), Web of Science (https://www.webofscience.com/), IEEE Xplore (Institute of Electrical and Electronics Engineers, https://ieeexplore.ieee.org/), Medline (Ovid, https://ovidsp.ovid.com/), CINAHL (EBSCO, https://www.ebsco.com/products/research-databases/cinahl-database), PsycINFO (Ovid, https://www.apa.org/pubs/databases/psycinfo), ArXiv (repository of preprints, https://arxiv.org/), Big Data Cognition and Computation (indexed journal, https://www.mdpi.com/journal/bdcc), PeerJ Computer Science (indexed journal, https://peerj.com/computer-science/), and Proceedings of international conferences organized by the ACM (Association for Computing Machinery, https://dl.acm.org/).

Search strategy: ((“artificial intelligence” OR “machine learning” OR algorithm OR “deep learning” OR “natural language processing” OR “AI-based” OR “AI-assisted”) AND (bias OR “algorithmic bias” OR fairness OR equity OR “social bias” OR “systematic bias” OR “reporting bias” OR “selection bias” OR “publication bias” OR “evaluation bias”) AND (“systematic review” OR “systematic reviews as topic” OR “evidence synthesis” OR “literature screening” OR “study selection” OR “abstract triage” OR (“abstract triage” OR (“systematic review” OR “systematic reviews” OR “abstract triage” OR “PRISM” OR “ROBIS” OR “meta-analysis” OR “automatic screening”) AND (health OR medicine OR “health sciences” OR clinical OR “primary health care” OR “electronic health records” OR “health informatics”)) AND ((taxonomies AND algorithmic bias stages) OR (detection and mitigation of bias) OR (transparency and reporting approaches) OR (visualization and illustration techniques) AND (guideline[Filter] OR meta-analysis[Filter] OR systematic review[Filter])).

Sources included were systematic reviews, conceptual frameworks, proposals for reporting guidelines, and empirical studies published in English or Spanish. The elements extracted included: (a) taxonomies by algorithmic bias stages; (b) strategies for the detection and mitigation of bias; (c) transparency and reporting approaches; and d) visualization and illustration techniques. The selection prioritized work that is comprehensive and clearly structured using breakdowns of pipeline stages, and those that reference or apply established reporting standards. Only the data and statements explicitly presented in the reference texts provided were included.

The search for articles closed on April 17, 2025.

The papers were selected and reviewed by the two authors.

The search and selection of articles was assisted with AI UnderMind.

Table 1 (Annex 1) shows the concepts that appear in this work.

**Table 1.** Key concepts explained for non-experts.

| Term | Definition |
| --- | --- |
| Adversarial learning | Training one model to do a task well while another tries to guess sensitive features |
| Algorithmic Bias | Systematic deviations introduced by AI algorithms that cause unfair results. Unfair or unequal decisions for certain groups |
| Artificial Intelligence, AI | A computer science discipline that simulates human cognitive processes. |
| Automatic Study Screening | AI-assisted process for filtering scientific articles. |
| Causal interventions | Identify and eliminate the root causes of bias |
| Checklist | Tool to ensure bias and transparency assessment. |
| Classical review biases | Types of biases in traditional systematic reviews (publication bias and research biases). |
| Cross-domain Examples | Illustrations of algorithmic biases in various contexts. |
| Demographic parity | Seek to ensure that the proportion of positive decisions is equal between different groups |
| Detection and Mitigation Strategies | Methods for identifying and reducing bias in AI systems. |
| Drift | Change in data distribution that affects model performance. |
| Equalized odds | Seek to ensure that the error rate is the same between different groups |
| Explainability | Ability to understand how and why a model makes a decision |
| Fairness Metrics | Indicators to measure the fairness of an algorithm. |
| Fairness-constrained optimization | Train the model to be accurate and fair |
| Fairness-performance trade-off | Conflict between improving fairness and maintaining accuracy |
| False Positive and False Negative Rate (FNR) Rates (FPR) | Proportion of cases misclassified as positive or negative. |
| Federated learning | Train models using data in different locations without moving it to a central server |
| Invariant learning | Identify consistent useful patterns in different situations |
| Model stitching | Combine parts of different models for accuracy and fairness |
| Modeling phase | Part of development where the model is trained |
| Pipeline-aware fairness | Consider equity at all stages of system development |
| Prejudice remover regularizer | Mathematical tools to reduce reliance on sensitive attributes |
| Preprocessing, In-processing, Post-processing | Adjustments made before, during, and after model training. |
| Reporting Standards | Guidelines and standards for documenting AI studies or models. |
| Reweighting techniques | Modifying the importance of certain examples during training |
| Sensitive attributes | Personal characteristics that should be treated with care |
| Shapley values | Calculate how much each characteristic influences a prediction |
| Spurious correlations | Casual or erroneous relationships between variables in the data |
| Systematic Review | A structured method for collecting and synthesizing evidence on a specific question. |
| Taxonomy | Hierarchical and systematic classification of elements or concepts. |
| Transfer learning | Reuse what a model learns in a previous task |
| Transferable representations | Intermediate ways to represent data that are unbiased |
| Transparency | Document and openly communicate all aspects of AI systems. |
| Visualization Techniques | Graphical methods for representing complex information. |
| Workflow | Sequence of phases in an AI project. |

## Results

The PRISMA flowchart in Figure 1 summarizes the selection of items.

**Figure 1.**
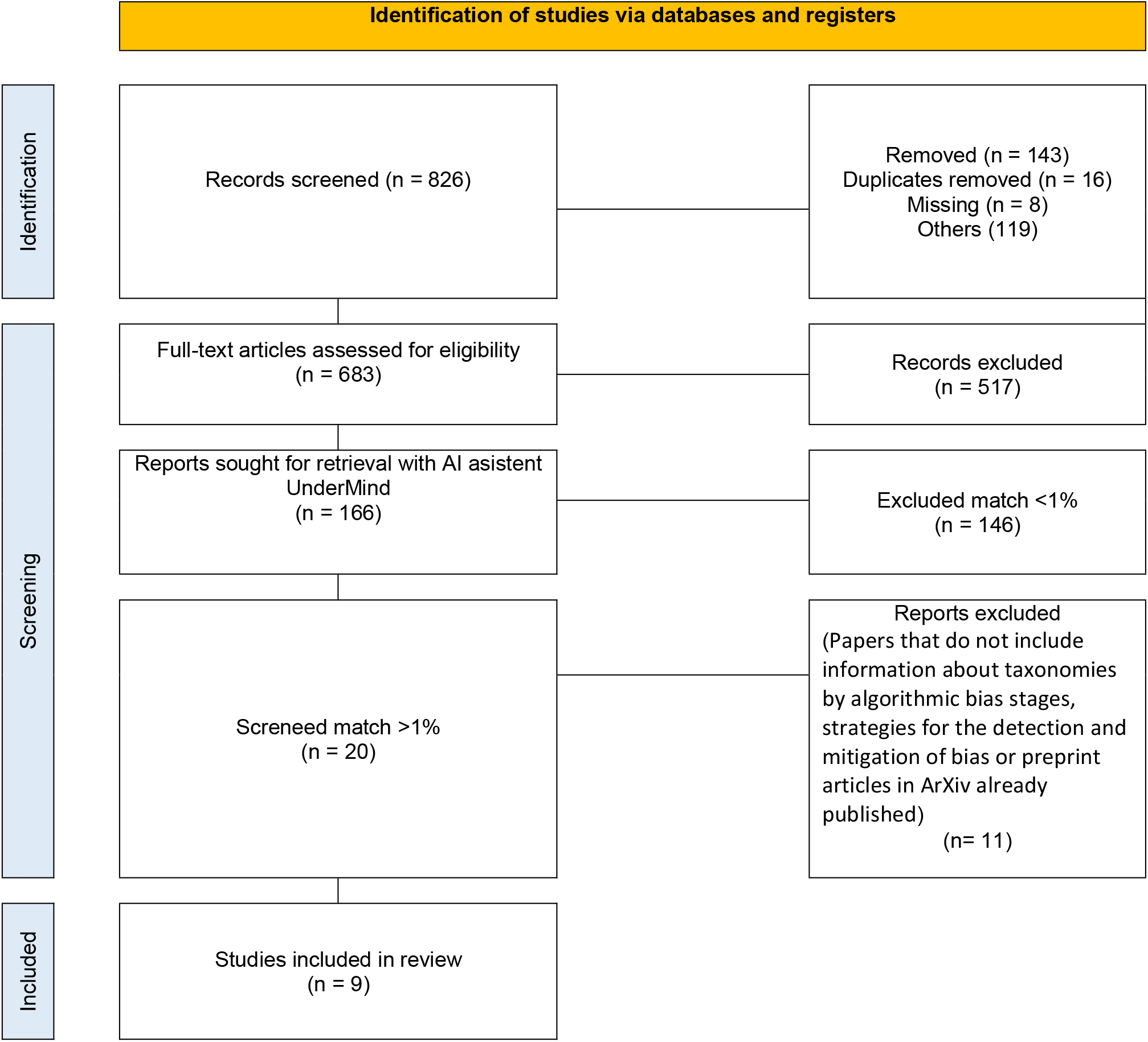
PRISMA 2020 flow diagram (Source: Page MJ, et al. BMJ 2021;372:n71. doi: 10.1136/bmj.n71) This work is licensed under CC BY 4.0. To view a copy of this license, visit https://creativecommons.org/licenses/by/4.0/

Out of 166 articles initially selected, 20 articles were included, from which 11 articles were finally excluded ^5–15^ - papers that do not include information about taxonomies by algorithmic bias stages, strategies for the detection and mitigation of bias or preprint articles in ArXiv already published - and 9 articles were included for the study ^1–4,16–20^.

### Staged taxonomies of algorithmic bias

Several comprehensive frameworks detail the categorization of algorithmic bias at each stage within health-related AI development processes as outlined in Table 2. Drukker et al. present a five-stage process (data collection, preparation/annotation, model development, evaluation, implementation) with 29 identified sources of bias, visually mapped through heat maps ^1^. Similar multiphase approaches are found in the reviews by Nazer et al., Chen et al., and Black et al., developing taxonomies that range from problem framing, through data acquisition and preprocessing, to implementation ^2,3,16,18,19^. These taxonomies provide explanations not accessible by experts and systematically assign types of bias to discrete stages of the pipeline. However, all the models analyzed focus on clinical prediction or imaging workflows; None apply directly to the context of systematic review selection workflows.

**Table 2.**
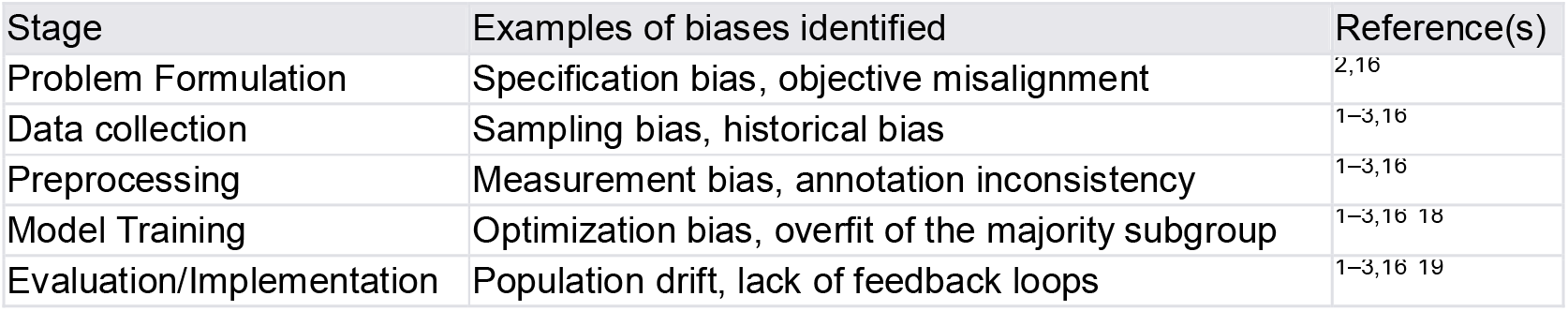
Example of a staged taxonomy of algorithmic bias in AI pipelines in healthcare.

| Stage | Examples of biases identified | Reference(s) |
| --- | --- | --- |
| Problem Formulation | Specification bias, objective misalignment | 2,16 |
| Data collection | Sampling bias, historical bias | 1-3,16 |
| Preprocessing | Measurement bias, annotation inconsistency | 1-3,16 |
| Model Training | Optimization bias, overfit of the majority subgroup | 1-3,16 18 |
| Evaluation/Implementation | Population drift, lack of feedback loops | 1-3,16 19 |

### Detection, mitigation, and reporting practices

Bias detection typically employs equity metrics such as statistical parity, equal opportunity, demographic parity, and stratified performance analysis (false positive/negative rates by subgroup) (Pagano et al. 2023; Chen et al. 2024). Mitigation strategies are grouped by pipeline phase: pre-processing (e.g., resampling, reweighting), in-processing (constraint optimization, adversarial learning), and post-processing (calibration, thresholds) ^2,3,18^ (see Table 3). Transparency recommendations include model versioning, script publishing, and using equity dashboards ^2,4,18^. Reporting standards such as CONSORT-AI and SPIRIT-AI are referenced as vehicles for documenting algorithmic bias, although there is no direct application within AI-assisted systematic reviews ^2,4^.

**Table 3.**
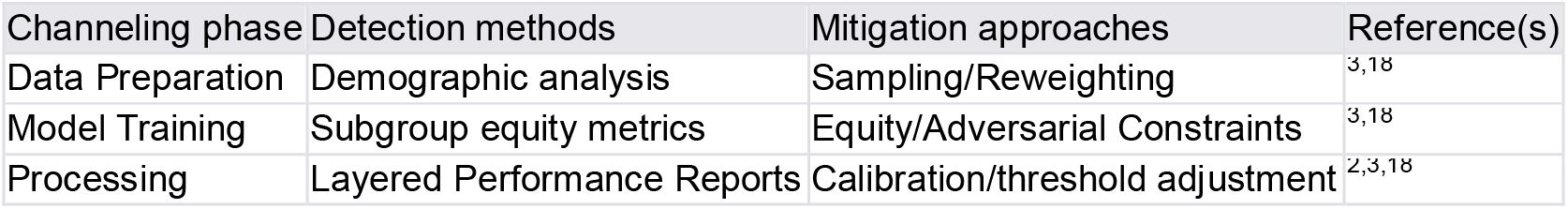
Bias detection and mitigation methods.

| Channeling phase | Detection methods | Mitigation approaches | Reference(s) |
| --- | --- | --- | --- |
| Data Preparation | Demographic analysis | Sampling/Reweighting | 3,18 |
| Model Training | Subgroup equity metrics | Equity/Adversarial Constraints | 3,18 |
| Processing | Layered Performance Reports | Calibration/threshold adjustment | 2,3,18 |

### Interactions with classic review biases

No identified studies empirically examine the interaction between algorithmic bias and classic systematic review biases (e.g., publication bias, selection bias, reporting bias) within AI-automated literature screening. While broad frameworks recognize that systemic, statistical, and human biases can coexist or amplify each other ^19^, no empirical or conceptual work contextualizes these interactions in systematic reviews.

### Visualization and Examples

Expert reviews and roadmaps employ visualizations, in particular, staged heatmaps, that demonstrate where risks of bias are concentrated in the AI development process ^1^. Equity curves, pipeline diagrams, and equity dashboards are recommended or described ^1–3,16 19^. Cross-domain examples (e.g., financial lending, malware detection) are used illustratively to explain the amplification of biases through AI, but without direct application to triage of the literature in the health sciences ^16,19^.

In the work of, Drukker et al. 2023, an example of a heat map is shown with the first column with the biases (one row for each type of bias), in the following columns the stages with the overview of the possible biases and where they are most likely to occur along the AI/ML pipeline of medical imaging, Dark shading with white dot indicates the most likely occurrence and lighter shading indicates additional potential occurrences (Figure 1, link to article: https://pubmed.ncbi.nlm.nih.gov/37125409/) ^1^.

## Discussion

The current literature demonstrates a mature landscape of algorithmic bias frameworks within the overall clinical development of AI. Multiple high-quality sources map the origins and consequences of bias at every step of AI pipelines, offer mitigation metrics and strategies, and propose greater transparency through standardized reporting ^1–3,16,18,19^. Strategies for practical mitigation, such as using de-bias algorithms at various stages of the pipeline and publishing model development details, are well described. However, none of the identified frameworks or studies address the assessment or reporting of algorithmic biases within AI-assisted screening for systematic reviews, nor do they empirically analyze how such biases interact with classic peer-level biases, such as publication or selection bias ^3,18,19^.

Visualization techniques, such as staged heatmaps and equity dashboards, are recognized as important for communicating risks of bias, but remain unused in the context of systematic review workflows. Recommendations for reporting and transparency (e.g., CONSORT-AI, SPIRIT-AI extensions, “ingredient” labeling), while robustly articulated for clinical prediction models, lack adaptation for systematic review processes that increasingly use AI-based screening ^2,4^.

The absence of direct assessment and reporting on algorithmic bias within the domain of AI-automated systematic reviews represents a critical and actionable gap. The adoption of existing taxonomies, reporting standards, and visualization techniques in this context can facilitate a more rigorous and equitable synthesis of evidence, particularly as AI continues to climb the lines of systematic review. The development of customized guidelines and empirical studies will be essential to close this translational gap and safeguard the integrity of automated literature reviews.

To conclude and as key points we will set some questions for future research, from a practical point of view it may be useful to specify the interest in evaluating algorithmic biases in systematic reviews together with the measurement of publication or research biases.

### How are algorithmic bias and research biases related in a systematic review

Both are different. Algorithmic bias: This mainly refers to systematic errors in automated decision systems (e.g., AI models) that produce unfair or unbalanced results for certain groups due to decisions in the design of the model, data, or the training process. Research biases (in systematic review): these are distortions that affect the validity of the findings of a review, including publication bias, understood as the tendency for studies with positive or significant results to be more likely to be published, selection bias when certain studies are systematically included while excluding others equally relevant, the language bias that occurs when only articles in certain languages are included, which filters the global picture or methodological biases due to the limited internal quality of primary studies (due to poor design, lack of control, etc.).

Table 4 summarizes the relationships between algorithmic bias and research biases.

**Table 4.** Relationship between algorithmic bias and research biases.

| Type of bias | How it relates to algorithmic bias in systematic reviews |
| --- | --- |
| Publication bias | It can fuel algorithmic bias: if automated systems (such as Undermind) are trained or adjusted using published literature, they replicate the patterns biased by this over-representation of positive outcomes. |
| Selection bias | If an algorithm prioritizes documents based on popularity (citations, impact), it can amplify selection bias if the most cited articles are not necessarily the fairest or methodologically sound. |
| Linguistic or geographic bias | If the automated system uses corpora in certain languages (e.g., English), it reproduces the cultural biases and region-centrism of those data in its results or document prioritization. |
| Biases in primary studies | Automatic search algorithms can give "valid" results of papers with non-obvious methodological errors, amplifying biased conclusions. |
| Algorithmic tool bias | The automated tool itself may have algorithmic biases during its modeling—for example, giving more weight to certain concepts or authors—which introduces a new level of specific bias to the review process. |

### Can the variability in responses between different AI applications or the same AI to the same research question be due to algorithmic bias?

Yes, it can be due to algorithmic bias for a variety of reasons.

If systematic variation in responses benefits or harms a specific group (for example: it always responds worse about certain languages, regions, disciplines, identities, or preferences), then we would be facing a case of algorithmic bias.

If the differences are purely random and not systematically correlated with sensitive attributes or other discriminating variables, then it would not be considered “algorithmic bias” in the strict sense, but rather randomness or stochastic variability.

Some research indicates that algorithmic bias can be amplified by specific modeling decisions, such as what type of architecture is used, how the model is trained, or which hyperparameters are chosen ^17,20^. That would explain why different systems (or even the same model in different settings) can offer uneven biased responses.

This variability may be due to generative AI models using techniques that incorporate randomness, such as seeds and stochastic sampling, as well as temperature adjustments during training, which generates different results for the same input. In addition, model design decisions, such as hyperparameters and random initialization, influence its final behavior. Differences between architectures and specific training objectives also result in different priorities in the types of responses they generate. Finally, asynchronous updates allow two instances of the same model to access different versions, affecting their knowledge base and behavior.

### Should failures in reproducibility be reported in the research report

Yes. If automated tools are used for review (such as generative AI, semantic search engines, etc.), an explicit discussion of potential algorithmic bias should be included as an additional source of bias in the systematic process.

This is directly supported by works such as Ganeesh et al. 2024, which document how different algorithmic decisions alter the system’s fairness metrics, which directly affects the neutrality of the literature search or prioritization ^17^. Bias mitigation techniques can vary in effectiveness depending on seemingly trivial elements, such as the order of training, pipeline configuration, or random seed. This suggests that technical variability should be considered as a methodological factor ^17^.

In rigorous research reports, transparency about reproducibility and potential sources of variability is critical, especially when using automated tools such as AI systems. The scientific precision in the methods allows the replication of the study, justifies possible margins of error when using AI, and recognizes the risk of systemic bias in the prioritization or ignorance of information.

### How to express it in a systematic review report

We offer some possible ideas to introduce in the writing of the research report of a systematic review.

### Formulation proposal in the method section

> *“An automated artificial intelligence system was used for the retrial and analysis of scientific literature. Importantly, due to the probabilistic and non-deterministic nature of the model, some variability was observed in the responses to identical queries. This variability may be due to internal modeling decisions, differences between versions of the system, or potential inherent algorithmic biases. Therefore, the results should be interpreted considering this possible lack of complete reproducibility. Multiple executions were performed to mitigate these effects and ensure robustness in the observed patterns*.*”*
>
> *“An artificial intelligence system analyzed scientific literature, and due to its probabilistic nature, the results varied between executions, suggesting the need to consider the lack of complete reproducibility when interpreting the observed patterns*.*”*
>
> *“The automated search system used is based on generative models with non-deterministic properties. As documented in recent studies, sensitivity to modeling choices such as training parameters or random seeds can impact model stability and fairness. To improve methodological transparency, this potential source of bias and variability has been documented as a limitation of the approach*.*”*
>
> *“Since part of the bibliographic selection was assisted by automated systems based on artificial intelligence, the possibility that these models introduced algorithmic biases was considered. In particular, algorithmic bias can interact with existing publication biases, amplifying the visibility of mostly positive studies or studies published in specific settings. Measures such as manual review of relevance and triangulation of sources were taken to mitigate these effects*.*”*
>
> *“To avoid algorithmic and publication bias in AI-assisted bibliographic selection, manual reviews and source triangulation were performed*.*”*

### b. Possible formulation in the limitations section

> *“An important limitation is related to the use of automated literature retrieval tools. In addition to the known presence of publication biases in academia, the system used could have introduced algorithmic bias during the prioritization and categorization of results. This dimension of bias – typical of system modeling – may have amplified certain types of studies or methodological approaches in a non-transparent way*.*”*
>
> *“Automated literature retrieval tools can introduce algorithmic bias, favoring certain studies or methodologies and amplifying publication bias*.*”*

## Conclusions

In a systematic review using artificial intelligence, algorithmic bias can amplify or filter publication bias.

Algorithmic bias poses an emerging threat to reproducible clinical evidence when automated tools are used.

Therefore, algorithmic bias can be considered an additional source of secondary bias. Simultaneous assessment of research, publication, and algorithmic bias is possible and necessary in systematic reviews.

Their inclusion in the methodology and limitations improves the transparency and quality of the review.

There are now comprehensive and staged frameworks for the identification and mitigation of algorithmic bias in health-related AI.

Reporting standards and visualization tools improve transparency in clinical AI.

However, the translation and operationalization of these methods in the workflow of AI-assisted systematic reviews, particularly in the study selection phase, remains unaddressed in the literature.

Research is needed to develop tailored guidelines, empirical assessments, and targeted visualizations to support equitable and bias-aware evidence synthesis in the health sciences.

New and standardized tools are required to objectively assess algorithmic bias in evidence-based medicine.

## Data Availability

All data produced in the present work are contained in the manuscript

## Annex 1

